# Autoantibodies to the *N*-Methyl-D-Aspartate Receptor in Adolescents with Early Onset Psychosis and Healthy Controls

**DOI:** 10.1101/2020.01.06.20016626

**Authors:** Kristine Engen, Laura Anne Wortinger, Kjetil Nordbø Jørgensen, Mathias Lundberg, Hannes Bohman, Runar Elle Smelror, Anne Margrethe Myhre, Leslie Jacobson, Angela Vincent, Ingrid Agartz

**Author notes:** **Correspondence** Dr. Laura Anne Wortinger, PhD.

## Abstract

**Background:** Autoantibodies to the *N*-methyl-D-aspartate receptor (NMDAR-Abs) in autoimmune encephalitis have been associated with prominent psychiatric symptoms. The aims of the present study are to identify the prevalence of NMDAR-Abs in adolescents with early onset psychosis disorders (EOP) and healthy controls (HC) and examine its clinical significance.

**Method:** Plasma samples were acquired from 46 adolescent EOP patients and 69 age- and sex matched HC, and assessed for the presence of immunoglobulin G NMDAR-Abs. All participants underwent psychiatric evaluation, neurological examination and head magnetic resonance imaging.

**Results:** NMDAR-Abs were detected in three of 46 (6.5%) EOP patients and in two of 69 (2.9%) HC. One NMDAR-Abs EOP patient presented with unusual psychopathology and minor T1 weighted lesions of vasculopathological origin located bi-frontally and in the basal ganglia, and had a recent diagnosis of a separate autoimmune disease. One NMDAR-Abs HC displayed a T2 weighted FLAIR hyperintensity lesion in the left frontal lobe. The remaining three NMDAR-Abs participants were two EOP patients without neurological or radiological findings, and one HC without any clinical findings.

**Conclusions:** We report the presence of NMDAR-Abs in both adolescent EOP patients and HC, similar to the incidence in other studies. This may support the hypothesis of an immunological disease component in a small proportion of adolescent psychosis, but the positive antibody tests must be carefully interpreted and reviewed within the individual clinical context.

## Introduction

Psychosis, as observed in severe mental illness, is a condition with unknown pathophysiology. The dopamine hypothesis only partially explains the symptoms occurring in affected individuals [1, 2], but pharmacological studies in both humans and animals indicate that hypoactivation of *N*-methyl-D-aspartate receptors (NMDAR) may cause the presynaptic hyperdopaminergia related to schizophrenia [3, 4]. Dysfunction in the glutamatergic system in patients with schizophrenia has been supported by positron-emission tomography (PET) and single-photon emission computed tomography (SPECT) studies, showing elevated levels of glutamate in the medial prefrontal cortex and the basal ganglia [5]. Within psychiatric research, the likelihood that a specific immune-mediated mechanism may define a biological subgroup of patients has gained considerable attention.

Anti-NMDA receptor encephalitis (ANRE) is an autoimmune neurological condition associated with IgG antibodies directed towards the NMDA receptor (NMDAR-Abs), which leads to NMDAR hypofunction [6]. There is sometimes a non-specific prodromal phase (viral-like illness) leading into psychiatric symptoms, most commonly delusions, auditory and visual hallucinations, mania, agitation, aggression, and unusual and bizarre behaviors [7]. Cognitive deficits and seizures with movement disorders can be observed within the first few weeks, with the possibility of hypoventilation and autonomic instability variably following [7]. Thus, ANRE has demonstrated further that loss of glutamatergic signaling can cause psychotic symptoms. Patients with ANRE are often seen initially by psychiatrists because of their psychiatric presentations [8] and indeed there are reports of ANRE patients with solely psychiatric symptoms, both in adult and in pediatric patients [9]. As a result, there have been many studies examining the prevalence of antibodies in adult psychiatric patients compared to healthy controls [10].

Early onset psychosis disorder (EOP) is defined as affective and non-affective psychotic disorders with disease onset before the age of 18 years [11]. EOP is a rare occurring condition with an estimated prevalence ranging from 0.05% to 0.5%, increasing during adolescence [12, 13]. EOP constitutes a group of severely debilitating conditions, including schizophrenia which is one of the main causes of disease burden in adolescents [14]. Within the EOP group, compared with adult onset patients, there are patients with poorer long term outcome [15], longer durations of untreated psychosis and a greater number of coexisting conditions [16]. To our knowledge, only one study on 43 pediatric EOP patients and 43 pediatric healthy controls assessed the presence of NMDAR-Abs and found an incidence of 5/43 (11.6%) in EOP without positives in the controls [17]. They found no clinical differences between antibody-positive and antibody-negative patients.

The study aim was to examine the prevalence of plasma IgG NMDAR-Abs in adolescent EOP patients and HC and to assess its association with age, sex, diagnosis and psychiatric features.

## Materials and Methods

### Participants

We examined the presence of NMDAR-Abs in 115 adolescents in this Scandinavian multi-center study with EOP (n = 46) and HC (n = 69). The study sample consisted of two clinical cohorts - one from the University of Oslo, Oslo, Norway and one from the Karolinska Institute, Stockholm, Sweden. Written informed consent was obtained from all participants and from parents/next-of-kin. The study was conducted in accordance with the Helsinki Declaration and approved by both the Swedish and the Norwegian National Committee for Ethics in Medical Research.

The Swedish sample consisted of 25 EOP patients and 34 HC participating in the Stockholm Child and Adolescent Psychosis Study (SCAPS). Patients aged 12-18 years of both sexes were recruited from the specialist care unit of psychosis and bipolar disorder in the department of Child and Adolescent Psychiatry at Sachs Children’s hospital in Stockholm, Sweden, and assessed by child- and adolescent psychiatry specialists working in the clinic. HC living in the Stockholm area were recruited using a population registry to ensure a similar demographic, age and sex distribution for the controls. The Norwegian sample consisted of 21 EOP patients and 35 HC participating in the Thematically Organized Psychosis research study for Youth (Youth-TOP), part of the Norwegian Centre for Mental Disorders Research (NORMENT). Patients aged 12-18 years of both sexes were recruited from the adolescent psychiatric inpatient units and outpatient clinics in the Oslo region and assessed by researchers consisting of trained psychiatrists and psychologists. HC, living in the Oslo area, were recruited from the Norwegian population registry, ensuring that the controls were similar to the included patients with regards to age and sex distribution and socio-demographic area. HC, for both samples, were excluded if they had been in contact with child and adolescent mental health care units, or if they currently met the criteria of a psychiatric disorder.

For both samples, inclusion criteria were (1) early onset psychosis according to DSM-IV (Diagnostic and Statistical Manual of Mental Disorders, 4^th^ edition), including schizophrenia spectrum disorders (schizophrenia, schizophreniform disorder, schizoaffective disorder), affective psychotic disorder (bipolar I disorder and major depressive disorder with psychotic features), and other psychotic disorders (psychotic disorder not otherwise specified (NOS), delusional disorder and brief psychotic disorder), (2) age between 12 and 18 years, (3) language abilities to complete interviews and self-rating tests and (4) written informed consent. General exclusion criteria were IQ < 70, previous moderate/severe head injury, a diagnosis of substance-induced psychotic disorder and organic brain disease.

### Diagnostic and Clinical Assessment with Regards to Psychiatric Diagnoses

The Swedish patients were assessed and diagnosed in accordance with ICD-10 [18] by their managing child and adolescent psychiatrists during their contact with the psychiatric unit. The Norwegian patients were diagnosed in accordance with the DSM-IV criteria using the Norwegian version of the Schedule for Affective Disorders and Schizophrenia for School Aged Children (6-18 years): Present and Lifetime Version (Kiddie-SADS) [19] and the Positive and Negative Syndrome Scale (PANSS) [20] for the assessment of current symptoms of psychosis. Functional outcome was assessed using the Children’s Global Assessment Scale (CGAS)[21], which is a measure of social, psychological and occupational functioning in both samples. Assessment included a general physical examination including a neurological examination consisting of the evaluation of gait, coordination, motor movements, reflexes and sensory examination. Data from the neurological examination was incomplete in the Swedish sample.

### Clinical Characterization and ANRE

Upon determining whether antibody positive participants had symptoms coinciding with ANRE; therefore, of particular interest in this study, we read through participants’ diagnostic and clinical assessments. These assessments were based on information from the participant, their parents and managing clinicians and from their medical records. Our goal was to identify possible symptoms and findings associated commonly with ANRE [8].

### Medication

Information regarding use of psychotropic medication in the Swedish sample was retrieved from the physicians treating the patients. For the Norwegian sample current and lifetime use of medication was collected from patients and/or their closest relatives using a structured questionnaire in an interview conducted by a research psychiatrist or psychologist. In some instances, information was retrieved from the physicians/psychologists treating the patients.

### NMDA Receptor Antibody (NMDAR-Abs) Measurements

Blood sampling was conducted at baseline for all participants. The plasma samples, which had been in a storage facility at −20 C, were sent to the Department of Clinical Neurosciences at the John Radcliffe Hospital in Oxford, England in 2015 for analysis of NMDAR-Abs.

NMDAR-Abs were measured (blind to the EOP or HC status of the sample) by indirect cell surface immunocytochemistry of transfected cells as used for routine diagnostic assays by the Oxford Neuroimmunology service since 2008. Briefly, live HEK293 cells are transfected with cDNAs of NR1 and NR2B subunits and incubated in human serum (1:20 initial dilution) or CSF (1:1 initial dilution) before fixation, followed by Alexa-Fluor labelled antihuman IgG. Antibodies binding to the cell surface of the live NMDAR-HEK293 cells were identified by fluorescence microscopy. A visual scoring system (0–4) is used and positive samples were repeated in a separate assay where they are also checked for specificity by lack of binding to HEK293 cells expressing an alternative antigen. As with other assays, scores of 0 are reported as negative and scores of 2–4 are reported as positive. Scores of 1 were initially reported as Low Positive, but after 2011 only scores of 1.5 are reported as Low Positive.

### MRI Acquisition and Assessment

Swedish participants underwent MRI scans on a 3 Tesla Discovery 750 scanner (GE Medical Systems) at the MR Research Centre at the Karolinska Institute in Stockholm. Norwegian participants were scanned using a 3 Tesla Signa HDxt scanner (GE Medical Systems) at the Department of Neuroradiology, Oslo University Hospital. A neuro-radiologist evaluated MRI images for pathological changes, including white matter hyper-intensity lesions. For both scanners, the assessment was based on structural MRI T1 weighted and T2-weighted fluid-attenuated inversion recovery (FLAIR) sequences.

## Results

Five of the 115 samples had detectable NMDAR-Abs, of which three of 46 (6.5%) were EOP patients and two of 69 (2.9%) were healthy participants.

Demographics and clinical features of antibody positive and antibody negative patients are shown in Table 1. The mean age of patients in the antibody positive and antibody negative groups was 14.5 ± 2.3 and 16.5 ± 1.8 years, respectively, with a significant difference being observed between groups (*p* = 0.026). There was a higher proportion of females in the study overall, with ratios of 72.1% and 66.7% in antibody-negative and antibody-positive groups, respectively. No significant difference in sex was observed between groups (*p* = 0.533).

**Table 1.** Results of statistical analysis.

|  | NMDA antibody positive patients (n=3) | NMDA antibody negative patients (n=43) | P-value | Statistical method |
| --- | --- | --- | --- | --- |
| Age, years (SD) | 14.46 ± 2.31 | 16.49 ± 1.17 | 0.026 | Fisher-Pitman permutation test |
| Sex (F/M) | 2/3 | 31/43 | 0.533 | Fisher's direct exact test |
| Diagnostic group (SZ/AFP/OTP) | 1/0/2 | 16/10/17 | 0.787 | Fisher's direct exact test |
| Age of onset, years (SD) | 13.81 ± 2.19 | 14.49 ± 2.17 | 0.508 | Fisher-Pitman permutation test |
| CGAS (SD) | 49.67 ± 12.86 | 41.35 ± 12.15 | 0.287 | Fischer-Pitman permutation test |

Abbreviations: SZ = schizophrenia spectrum disorders, AFP = affective psychotic disorders, OTP = other psychotic disorders. CGAS = children's global assessment scale

We found no significant difference in age of onset (*p* = 0.508). With regard to diagnostic group (schizophrenia spectrum disorders/affective psychotic disorders/other psychotic disorders), we found no significant difference between groups (*p* = 0.787). No significant difference in CGAS was observed between the groups (*p* = 0.287).

The five samples which were positive for NMDAR-Abs, had scores ranging from 1.5 (weak positive) to 2.0 (moderate positive). Clinical characteristic of the five participants are described in Table 2.

**Table 2.** Clinical features and MRI findings in NMDAR-Abs positive participants.

| Age and sex | Family psychiatric history | Presenting psychiatric symptoms | Somatic co-morbidity | Neurological symptoms | Head MRI | Psychiatric diagnosis | Treatment and outcome |
| --- | --- | --- | --- | --- | --- | --- | --- |
| 12, F | None | Affective lability. Aggression. Paranoid delusions. Suicidal ideation. | Juvenile rheumatoid arthritis | None | Multiple lesions of vasculopathological origin located bifrontally and in the right basal ganglia. | Psychotic disorder not otherwise specified 298.9 | Previously used aripiprazole for one year. Presently unmedicated. Several rounds of inpatient and outpatient treatment. |
| 16, M | Depression, father | Depressive symptoms. Depersonalization and derealization. Auditory, visual and tactile hallucinations. | None | None | Normal | Schizoaffective disorder, depressive type 295.7 | Previously tried sertraline and mirtazapine. Unmedicated during study. One previous inpatient admission. Ongoing outpatient treatment. |
| 14, F | Anxiety, maternal uncle | Anxiety. Depressive symptoms. Auditory hallucinations. Self harm. | None | None | Normal | Psychotic disorder not otherwise specified 298.9 | Previously used risperidone, aripiprazole and melatonin. Patient stopped taking medication with no increase in symptoms following this. Ongoing outpatient treatment. |
| 14, M | None | None | None | None | Normal | None | None |
| 17, F | None | None | None | None | 5 mm FLAIR hyper intensity lesion located in the left frontal lobe. Follow-up MRI taken 1 month later was normal (i.e. no lesion) | None | None |

Two of the three patients who tested positive for antibodies displayed typical psychiatric symptomatology and disease course with no neurological or radiological abnormalities. In these two patients, we found no evidence of the broad range of symptoms seen in ANRE, nor did we find any positive neurological signs or brain MR imaging findings such as white matter hyper intensities on T2-weighted FLAIR coinciding with ANRE. One of the NMDAR antibody positive HC had neither psychopathology, nor any neurological or MRI findings. We observed one patient and one control with NMDAR-Abs and ANRE related symptomatology or MRI findings.

### NMDAR-Abs Positive Adolescents with MRI Findings

The first participant of interest was a 12 year-old female who was diagnosed with psychotic disorder NOS. She had a prior diagnosis of obsessive-compulsive disorder (OCD) and body dysmorphic disorder (BDD), as well as juvenile rheumatoid arthritis (JRA), the latter being diagnosed months prior to the development of psychotic symptoms. Prior to illness onset, the participant exhibited good premorbid school and social functioning.

At the time of inclusion, she presented with paranoid delusions, affective lability, aggression and suicidal ideation as well as symptoms relating to OCD and BDD. Of note, there were no hallucinations, no apparent negative symptoms and no cognitive deficits noted on her neurocognitive examination. The neurological exam was without findings. MRI showed minor lesions of vasculopathological origin located bi-frontally and in the right basal ganglia (Figure 1). She was stabilized on aripiprazole with good effect on all of the symptoms she presented with at the time of inclusion. After two years, the antipsychotic medication was discontinued, and she has since been asymptomatic with regards to the psychotic disorder. A few months prior to starting antipsychotic medication, the patient began a successful treatment for JRA in the form of a tumor necrosis factor-alpha inhibitor.

**Figure 1:**
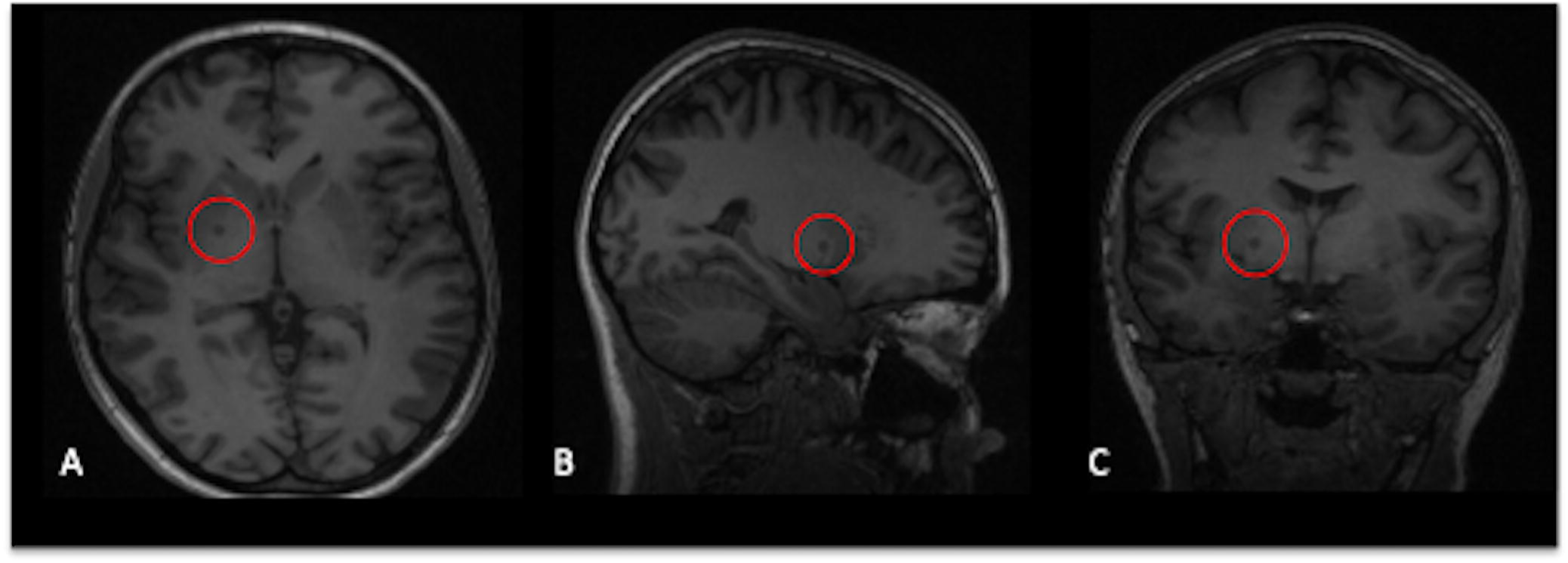
T1-weighted images showing several lesions of vasculopathological origin located bi-frontally and in the right basal ganglia, shown here in axial (a), sagittal (b) and coronal (c) view.

The second participant who tested positive for NMDAR-Abs was a 17 year-old female in the HC group. She had no prior or present psychopathology. The results from the neurocognitive assessment were within normal range on all tests, and she had no neurological findings upon inclusion. She had no prior somatic diseases or hospital admissions. She presented with a single finding on a FLAIR MR pulse sequence consisting of a 5 mm white matter hyper intensity lesion located in the left frontal lobe (Figure 2). This finding prompted a follow-up MRI, which was conducted one month later without observed abnormalities, i.e. the prior FLAIR abnormality was resolved. The same neuro-radiologist reviewed both scans. Neither at time of inclusion, nor at MRI follow-up scan, did she present with any symptoms or neurological findings coinciding with ANRE. Her school and social functioning remained unremarkable. Further clinical follow-up was not indicated.

**Figure 2:**
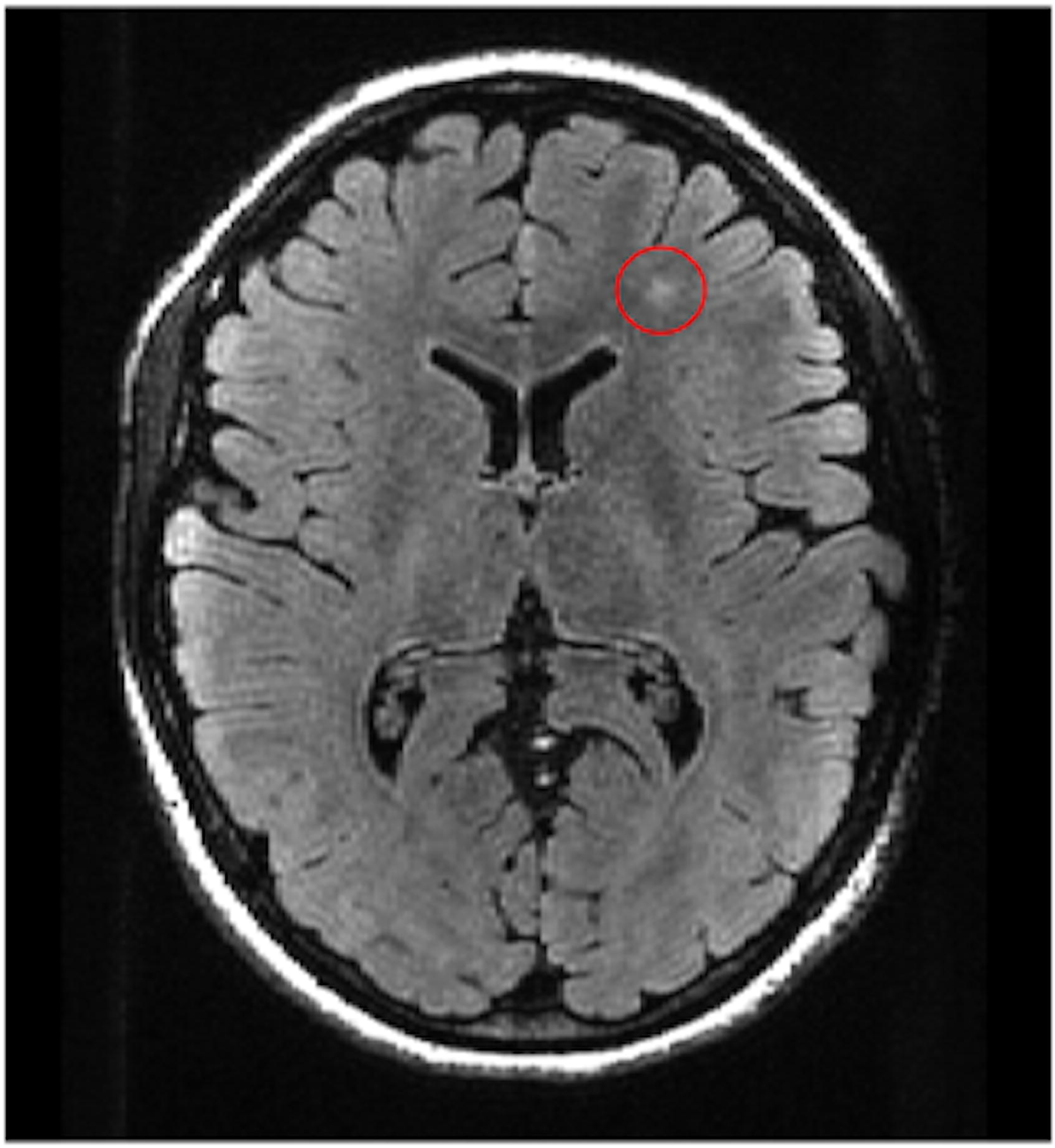
T2 weighted FLAIR image showing a 5 mm hyper intensity lesion in the left frontal lobe.

## Discussion

From the multi-center Scandinavian cohorts, we report three adolescents in the EOP patient group (6.5%) and two in the HC group (2.9%) who tested positive for NMDAR-Abs. Antibody-positivity was seen in both sexes, across psychiatric diagnostic groups, and did not affect function (measured by CGAS) compared to antibody negative EOP patients. Patients with antibodies were younger than patients without antibodies. Further assessment of antibody positive participants revealed a spectrum of clinical symptoms in the five participants and specific MR findings in two participants. An unusual psychiatric presentation was described in one of the participants.

Our finding of antibody-positive EOP patients is in line with Pathmanandavel and colleagues, the only previous study reported in an adolescent EOP patients [17]. They identified serum IgG NMDAR-Abs in five of the 43 EOP patients (11.6%). Patients in the Pathmanandavel et al., study were well characterized clinically, but neuroimaging was only performed in 38 of 43 patients, of which 27 had MRI head scans (eleven patients had computed tomography head scans). This is in contrast to our study, where all participants, both EOP patients and HC, had MRI head scans. They did not find antibodies in any of their 43 control participants. These were either healthy control subjects (n=17), patients with other neurological disease (epilepsy, cerebral palsy, neurometabolic disease, and neurodegenerative disorders), general medical control subjects (growth hormone deficiency, recurrent fractures, abdominal pain, infection), or inflammatory neurological control subjects (including first episode of demyelination). Their finding is in contrast to our results, as we identified HC with IgG antibodies. Though small sample sizes may explain the difference in these findings, other differences such as the use of carefully matched adolescent HC in our study, rather than partially pre-adolescent children (their controls ranged from 4-18 years of age and thus were younger than those in our study) and sensitivities and specificities of NMDAR-Abs assays used also could be an explanation.

The prevalence of NMDAR-Abs in our study is also consistent with studies on adult psychiatric patients, where a meta-analysis found that 21 of 1441 (1.46%) patients with schizophrenia and other psychotic disorders where positive for IgG NMDAR-Abs [10]. The same meta-analysis also identified that 5 of 1598 (0.3%) HC participants were IgG positive. Another study has shown that ANRE can have an atypical disease course consisting mainly of psychiatric symptoms [9]. In three antibody positive EOP patients identified in our study, two had psychosis without obvious neurological symptoms or findings, and unremarkable MRI scans. The third EOP patient with antibody-positivity was of particular interest due an unusual presentation of disease onset. This patient presented with a wide range of psychiatric symptoms, including comorbidity of OCD and BDD, abnormal findings bi-frontally and in the basal ganglia on brain MRI and a recent diagnosis of a juvenile rheumatoid arthritis. It is known that autoimmune diseases show a co-occurrence with other autoimmune diseases in as many as 25% of patients [22] perhaps partly due to shared genetic variants [23]. The antibody positive EOP patients in our study may represent a less severe disease course, rather than the full neurological presentation of ANRE.

One of the two controls with NMDAR-Abs had an MRI finding in the form of a 5 mm white matter hyperintensity lesion located in the left frontal lobe, which is the kind of hyperintensity lesion common in ANRE patients [8], but which had disappeared on subsequent scanning. White matter hyperintensity lesions are not uncommon in healthy populations, or in patients with psychotic disorders [24], but are most commonly seen during aging [25]. The significance of a fleeting MRI white matter intensity associated with a positive NMDAR-Abs test could be indicative of a transient form of ANRE in a susceptible age group, but may also be completely unrelated.

There are limitations. Several researchers in the field have pointed out difficulties in identification of the antibodies [26] and in particular how low antibody titers may be present in healthy individuals [10]. This is in line with the approach to autoimmune disorders in general; where low titers may not represent the manifestation of an autoimmune disorder, whilst higher titers are more likely to indicate the manifestation of a disease [27]. In addition, false positives are not uncommon in other autoimmune disorders [28]. This appears also to be the case with IgG NMDAR-Abs, as some studies have reported a seroprevalence of 1.2% and 0.4% in healthy populations [29, 30]. Moreover, we measured plasma antibodies rather than those in cerebrospinal fluid (CSF). A previous study on ANRE patients by Gresa-Arribas and colleagues showed a higher sensitivity for NMDAR-Abs in CSF rather than in serum (100% vs. 85.6%) [31].

Scientific research has shown that EOP patients have a less favorable prognosis compared to adult onset psychosis [15]. The number of studies conducted on EOP compared to adult psychotic disorders is fewer because of the low prevalence of EOP in a population. Our findings may represent a subgroup of patients with a range of symptoms associated with antibody-positivity. Studying adolescent EOP patients, it might be easier to identify the biological disease mechanisms as opposed to secondary phenomena associated with more long-term psychosis disease phase found in adults.

### Conclusions

We report the detection of IgG NMDAR-Abs in plasma in 6.5% of EOP patients and 2.9% of HC participants. Our findings may indicate that specific immune-mediated mechanisms define a biological subgroup of psychosis patients. However, the five antibody positive participants ranged from being symptom free to exhibiting symptoms and MRI findings coinciding with ANRE. We conclude that testing of patients must be done in an individual clinical context and that the presence of NMDAR-Abs must be carefully interpreted. Further studies conducted on independent samples of EOP adolescents are needed in order to increase understanding of the pathological significance of NMDAR-Abs in early psychosis.

## Data Availability

Data is available with some limitations.

## Acknowledgements

The authors would like to thank all participants and their families for their contribution to this study. We thank Dr. Vera Lonning for her important work in recruiting and assessing Norwegian EOP patients and controls. We express our gratitude to the funding agencies: the Research Council of Norway (#223273), the South-Eastern Norway Regional Health Authority (#2014-114), the Swedish Research Council (K2012-61X-15078-09-3, K2015-62X-15077-12-3 and 2017-00949) and FORMAS.

## Notes

### Competing Interest Statement

Angela Vincent and the University of Oxford hold patents for antibody assays, and Angela Vincent receives a proportion of royalties from Athena Diagnostics and Euroimmun AG. The remaining authors report no potential conflicts of interest.

### Funding Statement

IA was awarded funding from the Research Council of Norway, https://www.forskningsradet.no/en/ grant number 223273; the South-Eastern Norway Regional Health Authority https://www.helse-sorost.no/south-eastern-norway-regional-health-authority grant number 2014-114; the Swedish Research Council https://www.vr.se/english.html and FORMAS https://www.formas.se/en/ grant numbers K2012-61X-15078-09-3, K2015-62X-15077-12-3 and 2017-00949).

